# Trial protocol: RadTARGET, a multicenter phase II randomized controlled trial evaluating focal radiotherapy boost with de-intensification of dose to non-suspicious prostate in patients with intermediate- or high-risk prostate cancer

**DOI:** 10.64898/2026.04.18.26351182

**Authors:** Anna M Dornisch, Mariluz Rojo Domingo, Roberta Vezza Alexander, Christopher C Conlin, Son Do, Rana R McKay, Vitali Moiseenko, Michael A Liss, Jasmine Liu, Todd Pawlicki, Samuel Peña, Edmund M Qiao, Brent S Rose, Rhea Rupareliya, Ajay P Sandhu, Jessica Scholey, Steven N Seyedin, James J Urbanic, Lee-Jen Wei, Tyler M Seibert

**Author notes:** **Corresponding Author** Tyler M. Seibert, Mailing Address: ACTRI Building, 9500 Gilman Dr., MC 0861 La Jolla, CA 92093-0861. These authors contributed equally to this work (Anna M Dornisch, Mariluz Rojo Domingo). **Trial Information** ID: NCT06990542, Start Date: June 2025, Sponsor: University of California San Diego, Phase: Phase 2 Prostate Cancer Research Study, Study Type: Interventional.

## Abstract

Definitive radiotherapy (RT) for prostate cancer (PC) with dose intensification and/or focal boosting has excellent oncologic outcomes, but many patients experience adverse events. Dose escalation to the whole prostate improves outcomes at the expense of increased late adverse events. Intraprostatic recurrence after definitive RT typically occurs at the site of the primary tumor, suggesting that dose to the site of the dominant lesion is an important predictor of future failure. The efficacy and safety of tumor-focused RT compared to that of standard RT for definitive treatment of localized PC has not been assessed. RadTARGET (RAdiation Dose TAiloRing Guided by Enhanced Targeting) is a phase II randomized trial that aims to demonstrate superior safety of image-guided, tumor-focused RT compared to standard RT for acute genitourinary (GU) or gastrointestinal (GI) in the setting of definitive RT for intermediate- and high-risk PC. The study intervention is image-guided, tumor-focused RT with dose intensification of cancer visible on imaging and dose de-intensification to remaining prostate. Patients will be randomized to two arms: those who receive standard RT dose and those that receive tumor-focused RT. The study population will be patients with intermediate- or high-risk PC planning to undergo definitive RT with or without systemic therapy. The primary endpoint to compare between randomized arms is acute GU or GI grade ≥2 adverse events. Participant and study duration are 5 years and 8 years, respectively. RadTARGET will compare the efficacy and safety of tumor-focused RT to that of standard RT for definitive treatment of localized PC. We hypothesize that the tumor-focused approach will substantially reduce adverse events after prostate RT while retaining high efficacy. If this hypothesis is confirmed, we will conclude that a phase III randomized control trial is warranted to formally establish oncologic non-inferiority compared to the current standard of whole-gland dose escalation.

## INTRODUCTION and HYPOTHESES

Modern prostate radiation therapy (RT), with dose-escalation and focal boosting, has excellent oncologic outcomes for patients with intermediate- and high-risk prostate cancer (PC)^1,2^. Dose escalation to the whole prostate comes at the expense of meaningful adverse events^3,4^, spurring efforts to advance RT delivery to maximize both disease control and safety. The landmark FLAME randomized trial provided high-level evidence that the addition of a focal boost to the MRI-defined tumor improved oncologic outcomes without increased adverse events or decreased quality of life, compared to standard dose to the whole prostate^1^. However, many patients undergoing standard dose to the whole prostate still experience short- and long-term adverse events that impact post-treatment quality of life. Given that patients typically live many years after definitive treatment for localized PC, there is interest in improving the therapeutic ratio of definitive RT for localized PC to minimize impact of treatment on long-term quality of life.

One way to improve the therapeutic ratio is by decreasing rates of adverse events. Side effects of RT are driven by radiation dose to neighboring organs at risk (OARs), particularly rectum and bladder for definitive prostate radiotherapy^5^. The urethra and penile bulb are also thought to be important^6–8^. While focal boosting maintains a high probability of tumor control, treatment-related side effects could potentially be reduced through tumor-focused RT. Patterns of failure analyses in the context of both focal boosting and non-focal boosting demonstrate that intra-prostate recurrences occur primarily at the location of the dominant tumor^9–12^. Furthermore, the FLAME investigators demonstrated a clear dose-effect relationship: near-minimum dose to gross tumor volume (GTV) was associated with risk of PC recurrence^12,13^. Thus, intra-prostatic recurrence is likely a consequence of undertreatment of the primary tumor rather than undertreatment of the remaining prostate gland, providing a premise for a new approach to focal boosting with tumor-focused RT whereby the tumor receives ablative-dose RT, and the rest of the electively irradiated prostate receives a de-escalated dose.

Accurate discrimination of clinically significant intra-prostate tumors from benign prostate tissue is a prerequisite for tumor-focused RT. Multiparametric magnetic resonance imaging (mpMRI) is the current standard for PC detection^14^ and tumor delineation^15^. A quantitative imaging biomarker for PC – the Restriction Spectrum Imaging restriction score (RSIrs) – has been shown to be more specific than conventional mpMRI for clinically significant PC detection^16–19^. Beyond MRI, positron emission tomography targeting the prostate-specific membrane antigen (PSMA PET) is now routinely used in staging of primary PC^20^, and several studies report excellent performance for identification of intra-prostate tumors^21–23^. Furthermore, the combination of MRI and PSMA PET improve the sensitivity and specificity of intra-prostatic tumor delineation over each modality on its own^24,25^. By utilizing these advanced biological imaging techniques, intra-prostatic tumors can be accurately delineated with confidence, allowing for tumor-focused RT.

Tumor-focused RT involves ablative-dose RT to the imaging-defined tumors, while de-escalating dose to the remainder of the electively irradiated prostate gland. Similar approaches are commonly used in definitive RT for non-prostate malignancies, such as head and neck cancers. Lack of discrimination on CT scans between normal prostate tissue and PC previously made tumor-focused RT infeasible for PC. However, with improvement in imaging allowing for discrimination of PC versus benign prostate tissue, tumor-focused RT could now be adopted to lower dose to neighboring OARs and potentially increase safety and quality of life while preserving the excellent oncologic outcomes seen with current RT. Prospective data on this topic are scarce, and no randomized trials have been completed. There is one ongoing trial of MRI-guided RT dose de-escalation (DESTINATION) using a linear accelerator with onboard MRI (MR-linac) ^26^. A pragmatic, randomized trial with standard linear accelerators is warranted to compare the safety and efficacy of tumor-focused RT to standard RT. We describe here the RAdiotherapy Dose TAiloRing Guided by Enhanced Targeting (RadTARGET) study, a multicenter phase II randomized controlled trial evaluating a tumor-focused strategy for PC radiotherapy. We hypothesize that tumor-focused RT will yield similar efficacy and substantially lower adverse events compared to standard prostate RT.

### DESIGN

RadTARGET is a phase II, two-arm, randomized clinical trial evaluating tumor-focused RT versus standard-dose RT. Evaluation will be performed in terms of physician-reported adverse events, patient-reported safety, and oncologic outcomes. Participants will undergo RT for intermediate- or high-risk localized PC (Figure 1). A dynamic allocation procedure is used to assign participants in a 1:1 ratio to receive either standard-dose RT or image-guided, tumor-focused RT. To minimize biased clinical decision-making based on randomization, treating physicians will declare the participant’s treatment plan prior to randomization. Each arm will be balanced for risk group (intermediate/high), fractionation type (stereotactic body RT [SBRT] / non-SBRT), duration of androgen deprivation therapy (ADT; none/short-term/long-term), bladder filling (full/empty), and spacer use (yes/no). The primary objective is to evaluate the relative merits of image-guided, tumor-focused RT vs. standard-dose RT for acute GU/GI adverse events in the setting of definitive RT for intermediate- and high-risk PC. The endpoint will be acute, any-attribution GU/GI grade ≥2 adverse events within 3 months of completing RT, compared between randomized arms. Adverse events will be defined using Common Terminology Criteria for Adverse Events (CTCAE) version 5.0 grading.

**Figure 1.**
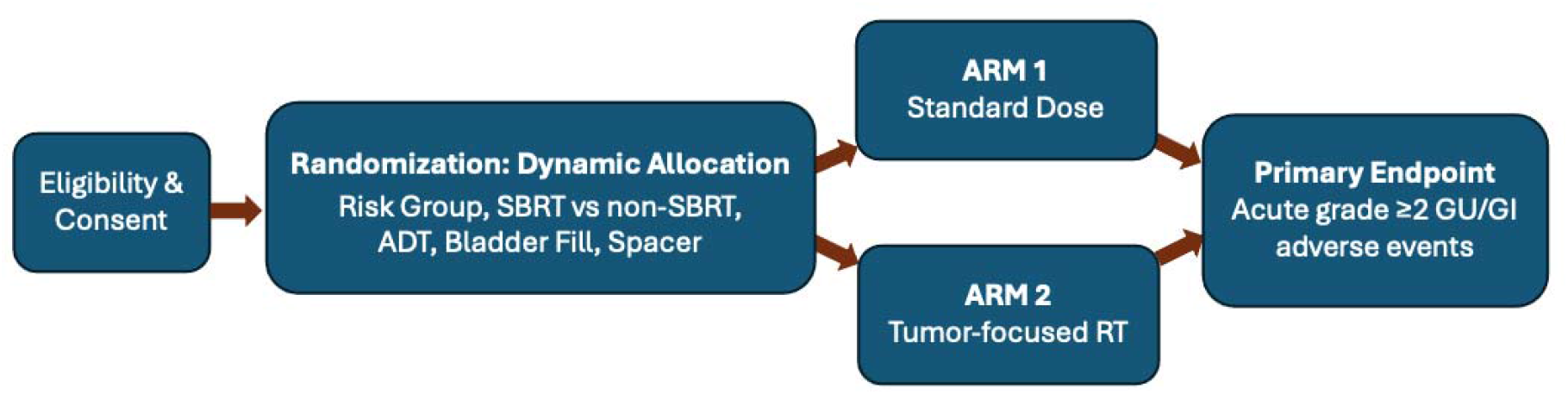
RadTARGET trial schema.

The study will enroll 150 participants, with completion of enrollment anticipated over 3 years. Participants must be ≥18 years old, have a diagnosis of intermediate- or high-risk localized PC, and plan to undergo curative-intent RT. They must also have one or more lesions visible on prostate MRI and/or PSMA PET with concordant pathology from biopsy needle locations. Participants with bilateral hip implants, prior PC treatment (including prostatectomy), or prior RT to the pelvis are not eligible. ADT may be started up to 90 days before randomization if study-compatible imaging was performed within 4 months of starting ADT. Participants must provide informed consent and be willing to comply with all study procedures for the duration of the study.

## PROTOCOL OVERVIEW

### Study Intervention

The study intervention is image-guided, ablative-dose RT to the sites of clinically significant, macroscopic tumors, while simultaneously de-escalating dose to areas of subclinical disease or benign prostate tissue within the rest of the electively irradiated prostate. All participants must have a pre-treatment MRI (consistent with PI-RADS v2.1 criteria^27^) with a multi-b-value diffusion-weighted MRI protocol compatible with calculation of the Restriction Spectrum Imaging restriction score (RSIrs) cancer biomarker. In RadTARGET, the treatment target (gross tumor volume [GTV]) will be manually delineated by the treating radiation oncologist using MRI with RSIrs +/- PSMA PET. Biopsy results and radiologist annotations that may be available should also be considered. For patients receiving ADT, the GTV should be delineated using pre-ADT images.

### Experimental Arm

For the experimental arm, clinical target volumes (CTV) are: *CTV_high*, defined as the GTV+5mm isotropic margin, cropped within prostate (with optional delineation of additional areas of moderate suspicion); and *CTV_low*, defined as the prostate +/- seminal vesicles (at the discretion of the treating physician). Elective pelvic lymph node RT (*CTV_LN*) is optional and at the discretion of the treating physician. Creation of planning target volumes (PTV) should follow UC San Diego Department of Radiation Medicine and Applied Sciences (RMAS) standards and guidelines, but adaptation is allowed at physician discretion or per local institutional standard practice. *PTV_high* is defined as *CTV_high* without additional margin. Doses for the various fractionations for the experimental arm are listed in Table 1. The dose covering ≥98% of the target volume (D98) for *CTV_high* for participants in the experimental arm must be at least standard of care prescription dose.

**Table 1.** Radiation doses for the experimental arm. Abbreviations: GTV, gross tumor volume (the visible tumor); CTV, clinical target volume (GTV with isotropic margin to cover areas at risk of microscopic disease); Gy, gray.

| Fractionation | GTV | CTV_high | CTV_low | CTV_LN |
| --- | --- | --- | --- | --- |
| **Standard Fractionation (39 – 40 fx)** | D98 up to 94 Gy | 78 – 80 Gy<br>in 39 – 40 fx | 56 Gy<br>in 28 fx | 50.4 Gy<br>in 28 fx |
| **Moderate Hypofractionation (28 fx)** | D98 up to 84 Gy | 70 Gy | 56 Gy | 50.4 Gy |
| **Moderate Hypofractionation (20 fx)** | D98 up to 74 Gy | 60 Gy | 56 Gy | 44 Gy |
| **Stereotactic Body Radiotherapy (5 fx)** | D98 up to 45 Gy | 40 Gy | 30 Gy | 25 Gy |

### Control Arm

As a pragmatic trial investigating image-guided, tumor-focused RT for intermediate- and high-risk prostate cancer, RadTARGET is not overly prescriptive for the control arm. Control arm definitive RT modality, dose, frequency, and administration must be in concordance with RMAS Standards and Guidelines or local institutional standard practice. Image-guided focal boost to MRI-visible disease is allowed and generally expected, but not required. For the control arm, the prostate CTV includes the entire prostate and receives the full prescription dose, corresponding to the relevant allowed fractionation scheme (i.e., *CTV_high* in Table 1). The proximal seminal vesicles may be included at physician discretion and would receive the *CTV_low* dose in Table 1.

### Both Arms

For both trial arms, all other CTV and PTV structures should conform to RMAS or local institutional standards and guidelines. OARs should be delineated per RMAS guidelines or local institutional practice. The urethra must be delineated if a focal tumor boost is delivered. The treating physician must state their intended radiation technique, number of fractions, coverage of lymph nodes, use of adaptive RT, and plan for spacer prior to randomization.

### Radiation Therapy Simulation (both arms)

Simulation allows for computer-based optimization of radiation dose delivery to the target tissue and simultaneous dose minimization to surrounding normal tissues. In both arms, simulation with CT and/or MRI is performed by RT-trained MR technologists per clinical routine. The diagnostic MRI +/- PSMA PET/CT are registered to the CT simulation for target delineation. A post-ADT planning MRI close to the date of the simulation is required for patients who began ADT more than 3 weeks prior to simulation. Full or empty bladder protocols are acceptable.

### Radiation Therapy Planning and Delivery (both arms)

RT planning is performed by trained dosimetrists and/or radiation physicists (complying with RMAS guidelines or local institutional practice) and documented in the medical records. Peer review of RT plans by radiation physicists and at least one radiation oncologist is required, as well as contouring of all OARs within 5 cm of any target volume. All plans must use modern planning and delivery techniques (e.g., VMAT), regardless of fractionation (conventional, hypofractionated, or SBRT). Target dose and/or coverage should be lowered to meet hard OAR dose goals, as needed. OAR dose goals may be exceeded in the patient’s interests at discretion of the treating physician. RT delivery is performed by trained RT therapists. A comparison of standard and tumor-focused plans for an example patient is shown in Figure 2.

**Figure 2.**
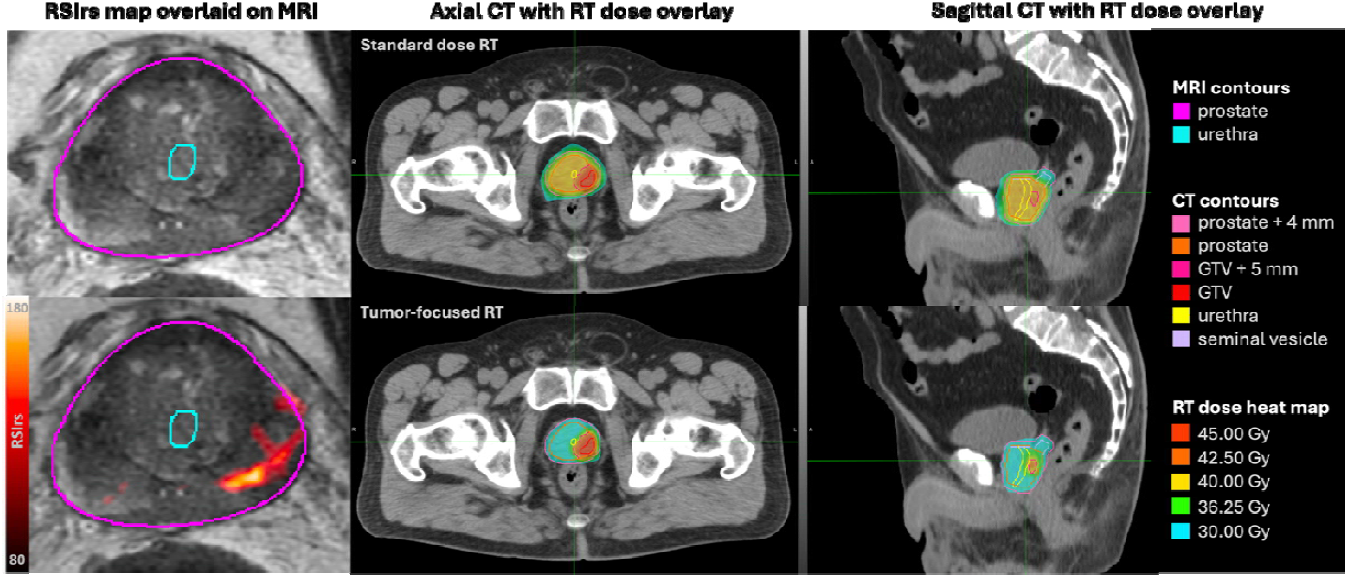
Comparison of standard dose and tumor-focused radiotherapy (RT) plans on computed tomography (CT) images for the RadTARGET trial. Magnetic resonance images (MRI) are used to localize the tumor for intensified treatment dose at the GTV (and for prostate and urethra contours). Restriction spectrum imaging restriction score (RSIrs) maps (overlaid on T2-weighted images) help radiation oncologists target the tumor. In the control arm, standard RT dose is applied to the entire prostate (with a uniform margin of 4 mm). Image-guided focal boost to MRI-visible disease is allowed and generally expected. In the experimental arm, the dose is increased at the tumor (with a uniform margin of 5 mm, limited to within the prostate) and decreased outside of the tumor. The patient shown was treated with Stereotactic Body Radiation Therapy (SBRT). Abbreviations: GTV=gross tumor volume, Gy=gray.

### Systemic Therapy

In both randomized arms, systemic therapy is given according to routine care. Use and duration of ADT is at the discretion of the treating physician. The intended use of ADT and/or other systemic therapy will be stated by the treating physician prior to randomization. Patients must also be on a stable dose of any medications to treat urinary symptoms, such as alpha-blockers of anticholinergic drugs, for at least 14 days prior to completion of baseline patient-reported outcomes questionnaires. After initiation of study protocol, medication to treat new urinary symptoms may be prescribed at discretion of treating physician (and will be recorded).

### Study Endpoints

- Primary Endpoint: physician-reported, all-attribution acute GI or GU grade ≥2 adverse events. Adverse events will be defined using CTCAE version 5.0 grading. This study will evaluate the relative merits of standard dose RT and image-guided tumor-focused RT in terms of safety, patient-reported outcomes, and oncologic efficacy.
- Secondary Endpoints:
  - Acute radiation-attribution GU/GI grade ≥2 adverse events.
  - Acute radiation-attribution grade ≥2 GU adverse events, other than alpha blockers.
  - Acute any-attribution and radiation-attribution grade ≥3 GU/GI adverse events.
  - Late any attribution and radiation-attribution grade ≥2 and grade ≥3 GU/GI adverse events.
  - Patient-reported urinary and bowel quality of life, defined as a decrease of 7 points and 6 points on the irritative/obstructive and bowel domains, respectively, of EPIC-26 at 3 months, 6 months, 12, 18, and 24 months after completion of RT.
  - Biochemical recurrence free survival at 5 years, defined as the percentage of participants alive and without biochemical at 5 years after completion of RT.
  - Local failure at 5 years, defined as the percentage of participants with intra- prostatic recurrence (as assessed by MRI and/or PSMA PET/CT or via confirmatory biopsy after biochemical recurrence) at 5 years after completion of RT.
  - Distant-metastasis-free survival at 5 years, defined as absence of local or regional lymph node recurrence (as confirmed on imaging including PSMA PET/CT and mpMRI) at 5 years after completion of RT.
  - Overall survival at 5 years, defined as the percentage of patients alive at 5 years after completion of RT.

### Data Collection and Follow-Up: Pre-therapeutic work-up and screening assessment

Screening evaluations must be performed within 42 days prior to randomization. Inclusion and exclusion criteria are checked and validated. The complete screening work-up includes medical history, medication review, demographic information, vital signs, performance status, and standard laboratory tests. Patients will report baseline quality of life assessment using EPIC-26, FACT-G, and prostate FACT questionnaires. When baseline assessments are completed, validation of inclusion and exclusion criteria for the study is performed followed by stratification parameter assessment. Patients are then randomized into tumor-focused RT or standard therapy arms.

### Data Collection and Follow-Up: Assessments during treatment phase

Patients undergo clinical assessments while undergoing RT per standard care clinical practice. On the last day of radiation treatment, patients undergo a study assessment. This will comprise of a medication review, vital signs, performance status, a provider adverse events assessment using the CTCAE v5.0 criteria, and a patient reported quality of life assessment using EPIC-26, FACT-G, and prostate-specific FACT (if applicable) questionnaires.

### Data Collection and Follow-Up: Assessments during the follow-up phase

Patients will have follow-up visits at 3, 6, 12, 18, and 24 months after randomization. At each follow-up visit, providers will assess medical history, medication review, performance status, and adverse events using CTCAE v5.0. This can be done via telehealth or in person. Additionally, patients undergo PSA +/- testosterone testing. Lastly, patients complete quality of life assessments using EPIC-26, FACT-G, and prostate-specific FACT questionnaires. Afterwards, long-term follow-up is conducted every 6 months (± 14 days) for at least 5 years after completion of RT to collect data on recurrence, metastasis, and survival. Long-term follow-up data can be extracted from the medical records. During the entire study, biochemical cancer recurrence is assessed by testing for PSA +/- testosterone level (testosterone measurement is optional during the long-term follow-up). Assessment of tumor recurrence and metastatic disease with MRI and/or PSMA PET or confirmatory biopsy after biochemical recurrence is done at the discretion of the treating physician.

### Data Collection and Follow-Up: Assessment of oncologic outcomes

Oncologic outcomes are assessed via PSA monitoring followed by routine clinical imaging and/or biopsy, if deemed appropriate at the discretion of the treating physician team. Biochemical failure is defined according to the Phoenix definition, and biochemical recurrence-free survival will be assessed. Rates of local failure will be assessed, and local failure will be defined as rate of intra-prostatic recurrence (as assessed on MRI and/or PSMA PET/CT or via confirmatory biopsy after biochemical recurrence). We will consider a local failure a primary site recurrence using the definition from the patterns of intra-prostatic failure analysis from FLAME with any overlap between the macroscopic recurrent tumor and the original *CTV_high*. A local failure will be a marginal miss if there is abutment between the macroscopic recurrent tumor and the original *CTV_high* without overlap. We will also assess distant metastasis-free survival (MFS) and regional + distant MFS (per FLAME secondary analysis), as confirmed on imaging, preferably PSMA PET/CT or mpMRI. Overall survival will also be assessed. Assessments will occur as per the Schedule of Activities (Table 2).

**Table 2.**
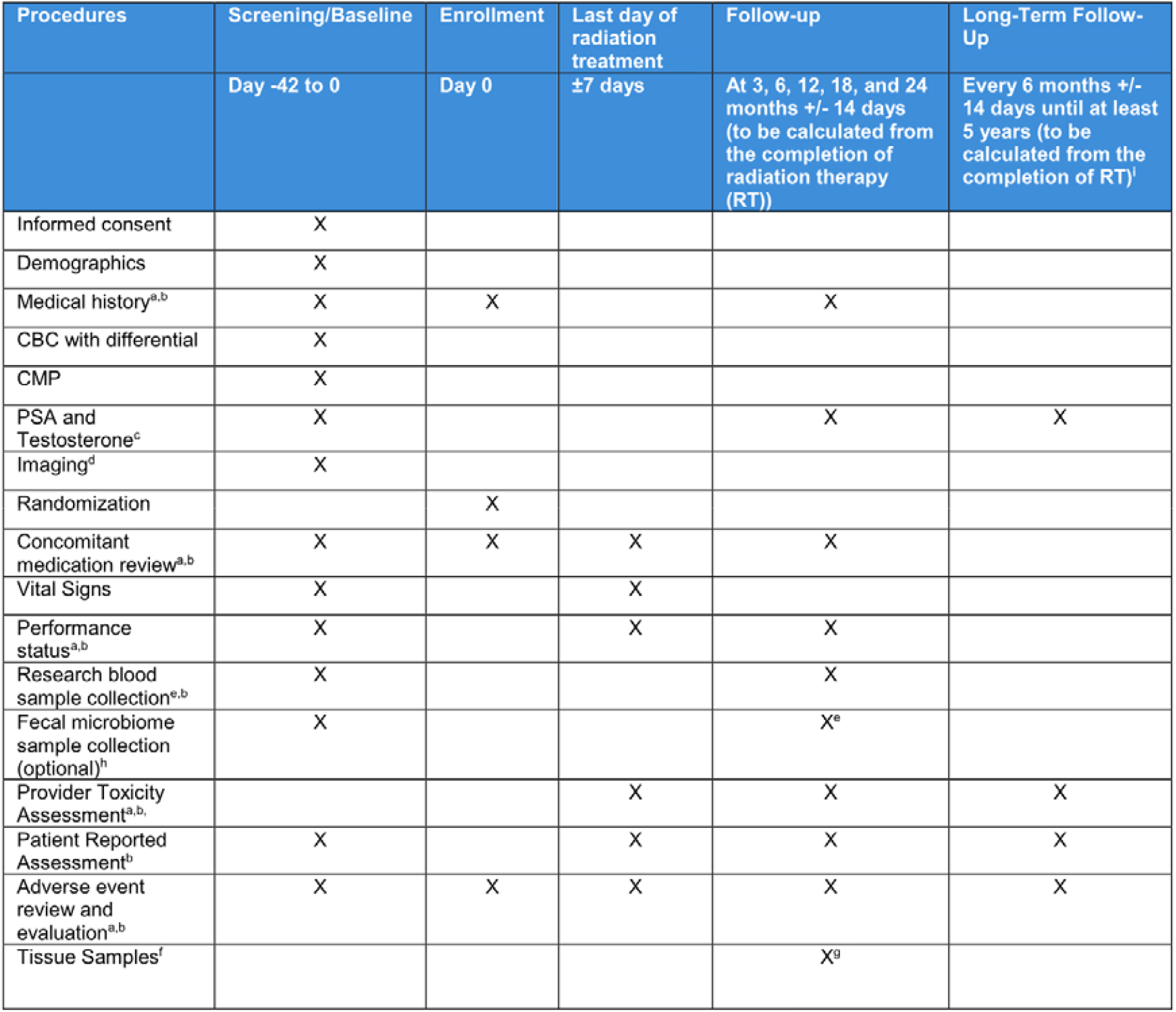
Schedule of activities. a: Via provider assessment, either telehealth or in person. b: After 12 months from completion of radiotherapy, it may be performed annually instead of every 6 months at investigator discretion. c: Testosterone measurement is optional for patients who do not undergo androgen deprivation therapy. Testosterone is also optional for all participants in long-term follow-up (i.e., >24 months after completing RT). d: Study- compatible MRI and (optional) PSMA PET/CT can be done within the 7 months prior to randomization. Hormone therapy may have begun within the 90 days prior to randomization, as long as study-compatible MRI and PSMA PET/CT were performed within the 4 months prior to starting the hormone therapy. It is preferred that pre-treatment imaging be performed within the 3 months prior to starting treatment. If hormone therapy precedes RT by ≥3 weeks, a new MRI must be obtained within 3 weeks of initiation of RT (preferably within 1 week). e: Research blood sample collection and optional stool sample collection will be performed at baseline and at 3-, 12-, and 24-months follow-up at time of standard of care lab draw. Baseline research blood samples can be collected any time before starting RT. f: From area of highest tumor content from prior biopsy or surgery: (a) 3 stained H&E slides and (b) 1 block or 30 unstained slides. g: Collection of archived tissue samples will be performed at an unscheduled visit. h: We invite study participants to optionally provide stool samples for exploratory microbiome analysis. i: Via chart review, phone call, telemedicine, or other means of secure communication with the participant to assess subsequent therapies and survival status. Continues until at least 5 years after completion of RT.

## STATISTICAL ANALYSIS

We adopt a statistical approach based on estimating the difference in adverse event rate (proportion of patients experiencing an adverse event) between the two randomized groups by controlling the precision (width) of the associated confidence interval for that difference. The primary endpoint is acute GU or GI grade ≥2 adverse events. We will estimate the absolute difference in adverse event rate between tumor-focused radiotherapy and standard dose radiotherapy using an intention-to-treat (ITT) analysis approach.

The following randomized trials and meta-analyses guide our assumptions for sample size determinations: RTOG 0415, PROFIT, HYPRO, PACE-B, FLAME, Hypo-FLAME, and MIRAGE^2,4,13,28–31^. From this high-level evidence, we assume the combined GU and/or GI acute adverse event rate is 40%. The MIRAGE trial included 156 participants and found a meaningful difference in acute GU and GI safety when comparing a difference in PTV margin size of 2 mm. We assume that the tumor-focused approach will lead to a similar adverse event reduction via the same mechanism.

We seek to answer whether the tumor-targeted approach reduces the rate of acute GU/GI adverse events. If we assume acute adverse events are reduced from 40% to 25% (absolute rate difference of -15%), we estimate that a sample size of 150 participants (75 in each arm) will yield a one-sided 90% confidence interval (one-sided equivalent to 95% confidence interval for a two-sided test) with 12.4% margin of error. Thus, we expect to have >95% confidence that the true rate difference is non-zero. We expect minimal attrition, but even 10% missing data would have a minimal impact on the margin of error (<1%). If adverse event rates in the standard arm were higher or lower than expected (30-70%), we would still have sufficient power to detect the assumed 15% absolute rate difference.

All primary and secondary analyses will be conducted with the intention-to-treat principle. All participants will be analyzed according to the group they were randomly assigned to, regardless of compliance with the trial protocol. Patients who did not receive RT treatment will be considered screen failures. The primary endpoints (proportion of patients with acute grade ≥2 GU or GI adverse events) will be compared across the two randomized groups using a stratified Cochran–Mantel–Haenszel test with the stratification factors listed above. We will also repeat the analysis using a non-stratified Fisher’s exact test.

Secondarily, we will develop a multivariable logistic regression model with binary dependent variable (presence or absence of acute grade ≥2 GU or GI adverse events) and independent variables to include randomization group and all covariates listed above. This will allow us to estimate the effects of each variable, with bootstrap 95% confidence intervals. The primary and secondary analyses above will be repeated in the following subgroups: (1) self-reported race/ethnicity, and (2) age <65 vs. ≥65 years. This is in addition to subgroup analyses within the stratification groupings for randomization.

### Translational Sub-studies

There is increasing interest in finding biomarkers predictive of cancer-directed therapy adverse events, including chemotherapy^32^ and RT^33^. Recently, radiogenomics studies have been identifying genetic markers that are associated with probably of treatment-related adverse events^34–37^. However, a recent systematic review found a large proportion of studies on correlation of genetic markers with radiotherapy-related side effects did not consider any dosimetric parameters^38^. There is a need for radiogenomics studies with complete information of dosimetric parameters as adverse events have been shown to be dose-dependent^39–41^. The focus of our biomarker investigation will include genomic analysis of peripheral blood samples for germline DNA, ctDNA, host immune cells, and tumor DNA. This allows for less invasive liquid biopsies that offer practical benefits including ease of collection and ability to measure tumor characteristics at multiple time points.

A genomic biomarker based on tumor expression of 22 genes (Decipher by Veracyte, South San Francisco, USA) is being evaluated as mechanism for treatment intensification/de-intensification in cooperative group trials for intermediate and high-risk prostate cancer. We will evaluate the Decipher score and its associated Decipher Genomics Resource for Intelligent Discovery (GRID) platform. There could be higher concern about the tumor-focused approach in patients with high-Decipher tumors. We will conduct subgroup analyses for those with low/intermediate (<0.60) vs. high (≥0.60) Decipher to learn whether a tumor-focused approach is more appropriate for tumors with lower (or higher) Decipher score. PORTOS is another score derived from the Decipher assay, with range -1 to +1, where higher scores have been shown to be associated with improved outcomes with dose-escalated radiotherapy^42^. However, higher PORTOS scores are also associated with worse genitourinary adverse events^43^. We hypothesize that those with higher PORTOS score (>0), indicating greater radiation response will favor a more focused approach and will have better outcomes in terms of reduced genitourinary adverse events.

Beyond liquid biopsies and tumor tissue, previous studies have reported that the microbiome may affect efficacy and safety of cancer therapy. A group of patient microbiomes was identified that may have a higher risk of gastrointestinal side effects during radiation therapy^44^. We invite trial participants to optionally provide samples for microbiome analysis and interested participants consent to sample collection.

## SUMMARY

RadTARGET is a phase II randomized trial designed to evaluate the safety and efficacy of a novel, tumor-focused approach to definitive RT for localized PC. The study leverages recent advances in imaging and RT delivery to individualize treatment plans for each patient with a goal of maintaining excellent efficacy while reducing adverse events compared to standard RT.

Importantly, we designed this trial with an eye toward improving treatment for most patients undergoing definitive RT for PC, not limited to patients with access to disease-site subspecialists at tertiary referral academic centers or with specialized, expensive equipment, such as an MR-Linac. Standard linear accelerators are used in RadTARGET. The FLAME patterns of recurrence analysis found that intra-prostatic recurrences overlapped with the GTV visible on pre-treatment MRI, underscoring that accurate delineation of MRI-visible tumors is critical for treatment success^13^. However, significant inter-observer variability in tumor contouring remains a challenge in clinical practice, especially when relying solely on conventional mpMRI^15,45^. While *important* in focal boosting, standardized, reproducible, and accurate tumor contours are imperative in tumor-focused RT to ensure consistency in treatment delivery and preservation of excellent oncologic outcomes.

For the GTV delineation in this trial, we mandate use of an advanced MRI biomarker (RSIrs), which provides superior tumor conspicuity, has been validated against histopathology in multiple studies, and is quantitative, allowing for standardized, reproducible thresholds in tumor identification^16–19^. While RSIrs is not widely available at present, it is a potentially scalable solution that can be implemented on modern clinical MRI scanners from any vendor. Thus, not only is RadTARGET one of the first randomized trials evaluating tumor-focused RT, but it is also the first randomized trial using a quantitative MRI biomarker to guide radiation target volumes in PC. Beyond MRI, our trial also allows PSMA PET/CT, if available, to complement MRI for GTV delineation, as the combination of MRI and PSMA PET improved the sensitivity and specificity of intra-prostatic tumor delineation over each modality on its own^24,25^. Our contouring strategy may be updated in the future if additional high-level data justify adjustments.

In conclusion, RadTARGET is a multi-center phase II randomized controlled trial evaluating an innovative, tumor-focused RT strategy for patients with intermediate- or high-risk PC. We hypothesize that the tumor-focused approach will substantially reduce adverse events after prostate RT while retaining high efficacy. If this hypothesis is confirmed, we will conclude that a phase III randomized controlled trial is warranted to formally establish oncologic non-inferiority compared to the current standard of whole-gland dose escalation.

## Data Availability

All data produced in the present study are available upon reasonable request to the authors

## Acknowledgements

This work was funded by UC San Diego School of Medicine and UC San Diego Moores Cancer Center.

## References

1. Kerkmeijer LGW, Groen VH, Pos FJ, et al. Focal Boost to the Intraprostatic Tumor in External Beam Radiotherapy for Patients With Localized Prostate Cancer: Results From the FLAME Randomized Phase III Trial. J Clin Oncol. 2021;39(7):787–796. doi:10.1200/JCO.20.02873

2. van As N, Griffin C, Tree A, et al. Phase 3 Trial of Stereotactic Body Radiotherapy in Localized Prostate Cancer. N Engl J Med. 2024;391(15):1413–1425. doi:10.1056/NEJMoa2403365

3. Pasalic D, Kuban DA, Allen PK, et al. Dose Escalation for Prostate Adenocarcinoma: A Long-Term Update on the Outcomes of a Phase 3, Single Institution Randomized Clinical Trial. Int J Radiat Oncol Biol Phys. 2019;104(4):790–797. doi:10.1016/j.ijrobp.2019.02.045

4. Michalski JM, Moughan J, Purdy J, et al. Effect of Standard vs Dose-Escalated Radiation Therapy for Patients With Intermediate-Risk Prostate Cancer: The NRG Oncology RTOG 0126 Randomized Clinical Trial. JAMA Oncol. 2018;4(6):e180039. doi:10.1001/jamaoncol.2018.0039

5. Olsson CE, Jackson A, Deasy JO, Thor M. A systematic post-QUANTEC review of tolerance doses for prostate cancer radiotherapy. International journal of radiation oncology, biology, physics. 2018;102(5):1514. doi:10.1016/j.ijrobp.2018.08.015

6. Groen VH, van Schie M, Zuithoff NPA, et al. Urethral and bladder dose–effect relations for late genitourinary toxicity following external beam radiotherapy for prostate cancer in the FLAME trial. Radiotherapy and Oncology. 2022;167:127–132. doi:10.1016/j.radonc.2021.12.027

7. Song Y, Nguyen L, Dornisch AM, et al. Urethra contours on MRI: Multidisciplinary consensus educational atlas and reference standard for artificial intelligence benchmarking. Radiotherapy and Oncology. 2026;214. doi:10.1016/j.radonc.2025.111231

8. Achard V, Zilli T, Lamanna G, et al. Urethra-Sparing Prostate Cancer Stereotactic Body Radiation Therapy: Sexual Function and Radiation Dose to the Penile Bulb, the Crura, and the Internal Pudendal Arteries From a Randomized Phase 2 Trial. International Journal of Radiation Oncology*Biology*Physics. 2024;119(4):1137–1146. doi:10.1016/j.ijrobp.2023.12.037

9. Pucar D, Hricak H, Shukla-Dave A, et al. Clinically Significant Prostate Cancer Local Recurrence After Radiation Therapy Occurs at the Site of Primary Tumor: Magnetic Resonance Imaging and Step-Section Pathology Evidence. International Journal of Radiation Oncology, Biology, Physics. 2007;69(1):62–69. doi:10.1016/j.ijrobp.2007.03.065

10. Arrayeh E, Westphalen AC, Kurhanewicz J, et al. Does Local Recurrence of Prostate Cancer After Radiation Therapy Occur at the Site of Primary Tumor? Results of a Longitudinal MRI and MRSI Study. International Journal of Radiation Oncology, Biology, Physics. 2012;82(5):e787–e793. doi:10.1016/j.ijrobp.2011.11.030

11. Cellini N, Morganti AG, Mattiucci GC, et al. Analysis of intraprostatic failures in patients treated with hormonal therapy and radiotherapy: implications for conformal therapy planning. International Journal of Radiation Oncology, Biology, Physics. 2002;53(3):595–599. doi:10.1016/S0360-3016(02)02795-5

12. Menne Guricová K, Pos FJ, Schoots IG, et al. Intra-prostatic recurrences after radiotherapy with focal boost: Location and dose mapping in the FLAME trial. Radiother Oncol. 2024;201:110535. doi:10.1016/j.radonc.2024.110535

13. Groen VH, Haustermans K, Pos FJ, et al. Patterns of Failure Following External Beam Radiotherapy With or Without an Additional Focal Boost in the Randomized Controlled FLAME Trial for Localized Prostate Cancer. Eur Urol. 2022;82(3):252–257. doi:10.1016/j.eururo.2021.12.012

14. Turkbey B, Rosenkrantz AB, Haider MA, et al. Prostate Imaging Reporting and Data System Version 2.1: 2019 Update of Prostate Imaging Reporting and Data System Version 2. Eur Urol. 2019;76(3):340–351. doi:10.1016/j.eururo.2019.02.033

15. Dornisch A, Zhong A, Poon D, Tree A, Seibert T. Focal radiotherapy boost to MR-visible tumor for prostate cancer: a systematic review. World J Urol. doi:10.1007/s00345-023-04745-w

16. Conlin CC, Feng CH, Rodriguez-Soto AE, et al. Improved Characterization of Diffusion in Normal and Cancerous Prostate Tissue Through Optimization of Multicompartmental Signal Models. J Magn Reson Imaging. 2021;53(2):628–639. doi:10.1002/jmri.27393

17. Feng CH, Conlin CC, Batra K, et al. Voxel-level Classification of Prostate Cancer on Magnetic Resonance Imaging: Improving Accuracy Using Four-Compartment Restriction Spectrum Imaging. J Magn Reson Imaging. 2021;54(3):975–984. doi:10.1002/jmri.27623

18. Domingo MR, Do DD, Conlin CC, et al. Restriction Spectrum Imaging as a quantitative biomarker for prostate cancer with reliable positive predictive value. medRxiv. Preprint posted online June 6, 2024:2024.06.05.24308468. doi:10.1101/2024.06.05.24308468

19. Do DD, Domingo MR, Conlin CC, et al. Robustness of a Restriction Spectrum Imaging (RSI) quantitative MRI biomarker for prostate cancer: assessing for systematic bias due to age, race, ethnicity, prostate volume, medication use, or imaging acquisition parameters. medRxiv. Preprint posted online September 12, 2024:2024.09.10.24313042. doi:10.1101/2024.09.10.24313042

20. Hofman MS, Lawrentschuk N, Francis RJ, et al. Prostate-specific membrane antigen PET-CT in patients with high-risk prostate cancer before curative-intent surgery or radiotherapy (proPSMA): a prospective, randomised, multicentre study. The Lancet. 2020;395(10231):1208–1216. doi:10.1016/S0140-6736(20)30314-7

21. Draulans C, De Roover R, van der Heide UA, et al. Optimal 68Ga-PSMA and 18F-PSMA PET window levelling for gross tumour volume delineation in primary prostate cancer. Eur J Nucl Med Mol Imaging. 2021;48(4):1211–1218. doi:10.1007/s00259-020-05059-4

22. Spohn S, Jaegle C, Fassbender TF, et al. Intraindividual comparison between 68Ga-PSMA-PET/CT and mpMRI for intraprostatic tumor delineation in patients with primary prostate cancer: a retrospective analysis in 101 patients. Eur J Nucl Med Mol Imaging. 2020;47(12):2796–2803. doi:10.1007/s00259-020-04827-6

23. Zamboglou C, Spohn SKB, Adebahr S, et al. PSMA-PET/MRI-Based Focal Dose Escalation in Patients with Primary Prostate Cancer Treated with Stereotactic Body Radiation Therapy (HypoFocal-SBRT): Study Protocol of a Randomized, Multicentric Phase III Trial. Cancers. 2021;13(22):5795. doi:10.3390/cancers13225795

24. Bettermann AS, Zamboglou C, Kiefer S, et al. [68Ga-]PSMA-11 PET/CT and multiparametric MRI for gross tumor volume delineation in a slice by slice analysis with whole mount histopathology as a reference standard – Implications for focal radiotherapy planning in primary prostate cancer. Radiotherapy and Oncology. 2019;141:214–219. doi:10.1016/j.radonc.2019.07.005

25. Eiber M, Weirich G, Holzapfel K, et al. Simultaneous 68Ga-PSMA HBED-CC PET/MRI Improves the Localization of Primary Prostate Cancer. European Urology. 2016;70(5):829–836. doi:10.1016/j.eururo.2015.12.053

26. Royal Marsden NHS Foundation Trust. A Feasibility Study of Dose De-Escalation in Prostate Radiotherapy Using the Magnetic Resonance Linear Accelerator (MRL). clinicaltrials.gov; 2024. Accessed October 22, 2025. https://clinicaltrials.gov/study/NCT05709496

27. Turkbey B, Rosenkrantz AB, Haider MA, et al. Prostate Imaging Reporting and Data System Version 2.1: 2019 Update of Prostate Imaging Reporting and Data System Version 2. Eur Urol. 2019;76(3):340–351. doi:10.1016/j.eururo.2019.02.033

28. Sanmamed N, Dayes I, Catton C, et al. Patient-reported Quality of Life in PROFIT, a Phase 3 Randomized Clinical Trial Evaluating Moderately Hypofractionated Radiotherapy for Intermediate-risk Prostate Cancer. European Urology Oncology. 2026;9(1):26–36. doi:10.1016/j.euo.2025.03.017

29. Incrocci L, Wortel RC, Alemayehu WG, et al. Hypofractionated versus conventionally fractionated radiotherapy for patients with localised prostate cancer (HYPRO): final efficacy results from a randomised, multicentre, open-label, phase 3 trial. Lancet Oncol. 2016;17(8):1061–1069. doi:10.1016/S1470-2045(16)30070-5

30. Draulans C, Haustermans K, Pos FJ, et al. Stereotactic body radiotherapy with a focal boost to the intraprostatic tumor for intermediate and high risk prostate cancer: 5-year efficacy and toxicity in the hypo-FLAME trial. Radiotherapy and Oncology. 2024;201. doi:10.1016/j.radonc.2024.110568

31. Kishan AU, Ma TM, Lamb JM, et al. Magnetic Resonance Imaging-Guided vs Computed Tomography-Guided Stereotactic Body Radiotherapy for Prostate Cancer: The MIRAGE Randomized Clinical Trial. JAMA Oncol. 2023;9(3):365–373. doi:10.1001/jamaoncol.2022.6558

32. Boguszewicz Ł. Predictive Biomarkers for Response and Toxicity of Induction Chemotherapy in Head and Neck Cancers. Front Oncol. 2022;12:900903. doi:10.3389/fonc.2022.900903

33. Rosenstein BS. Radiogenomics: Identification of Genomic Predictors for Radiation Toxicity. Semin Radiat Oncol. 2017;27(4):300–309. doi:10.1016/j.semradonc.2017.04.005

34. Alsner J, Andreassen CN, Overgaard J. Genetic Markers for Prediction of Normal Tissue Toxicity After Radiotherapy. Seminars in Radiation Oncology. 2008;18(2):126–135. doi:10.1016/j.semradonc.2007.10.004

35. Cesaretti JA, Stock RG, Lehrer S, et al. ATM sequence variants are predictive of adverse radiotherapy response among patients treated for prostate cancer. International Journal of Radiation Oncology*Biology*Physics. 2005;61(1):196–202. doi:10.1016/j.ijrobp.2004.09.031

36. Cesaretti JA, Stock RG, Atencio DP, et al. A Genetically Determined Dose–Volume Histogram Predicts for Rectal Bleeding among Patients Treated With Prostate Brachytherapy. International Journal of Radiation Oncology*Biology*Physics. 2007;68(5):1410–1416. doi:10.1016/j.ijrobp.2007.02.052

37. Yuan X, Liao Z, Liu Z, et al. Single nucleotide polymorphism at rs1982073:T869C of the TGFbeta 1 gene is associated with the risk of radiation pneumonitis in patients with non-small-cell lung cancer treated with definitive radiotherapy. J Clin Oncol. 2009;27(20):3370–3378. doi:10.1200/jco.2008.20.6763

38. Yahya N, Chua XJ, Manan HA, Ismail F. Inclusion of dosimetric data as covariates in toxicity-related radiogenomic studies. Strahlenther Onkol. 2018;194(8):780–786. doi:10.1007/s00066-018-1303-5

39. Matsuo Y, Shibuya K, Nakamura M, et al. Dose--volume metrics associated with radiation pneumonitis after stereotactic body radiation therapy for lung cancer. Int J Radiat Oncol Biol Phys. 2012;83(4):e545–9. doi:10.1016/j.ijrobp.2012.01.018

40. Yahya N, Ebert MA, Bulsara M, et al. Urinary symptoms following external beam radiotherapy of the prostate: Dose–symptom correlates with multiple-event and event-count models. Radiotherapy and Oncology. 2015;117(2):277–282. doi:10.1016/j.radonc.2015.10.003

41. Yoon H, Oh D, Park HC, et al. Predictive factors for gastroduodenal toxicity based on endoscopy following radiotherapy in patients with hepatocellular carcinoma. Strahlenther Onkol. 2013;189(7):541–546. doi:10.1007/s00066-013-0343-0

42. Dal Pra A, Ghadjar P, Ryu HM, et al. Predicting dose response to prostate cancer radiotherapy: validation of a radiation signature in the randomized phase III NRG/RTOG 0126 and SAKK 09/10 trials. Ann Oncol. 2025;36(5):572–582. doi:10.1016/j.annonc.2025.01.017

43. Hoffman KE, Kamran SC, Ryu HM, et al. PORTOS gene signature as a predictor of risk of adverse events after dose-escalated vs. lower-dose prostate radiation therapy in NRG/RTOG 0126. JCO. 2025;43(5_suppl):375–375. doi:10.1200/JCO.2025.43.5_suppl.375

44. Madkour MA, Altaf RA, Sayed ZS, et al. The Role of Gut Microbiota in Modulating Cancer Therapy Efficacy. Advanced Gut & Microbiome Research. 2024;2024(1):9919868. doi:10.1155/2024/9919868

45. Lui AJ, Kallis K, Zhong AY, et al. ReIGNITE Radiation Therapy Boost: A Prospective, International Study of Radiation Oncologists’ Accuracy in Contouring Prostate Tumors for Focal Radiation Therapy Boost on Conventional Magnetic Resonance Imaging Alone or With Assistance of Restriction Spectrum Imaging. Int J Radiat Oncol Biol Phys. 2023;117(5):1145–1152. doi:10.1016/j.ijrobp.2023.07.004

